# A phase 2A trial of the safety and tolerability of increased dose rifampicin and adjunctive linezolid, with or without aspirin, for HIV-associated tuberculous meningitis (The LASER-TBM Trial)

**DOI:** 10.1101/2022.07.26.22278065

**Authors:** Angharad G Davis, Sean Wasserman, Cari Stek, Mpumi Maxebengula, C. Jason Liang, Stephani Stegmann, Sonya Koekemoer, Amanda Jackson, Yakub Kadernani, Marise Bremer, Remy Daroowala, Saalikha Aziz, Rene Goliath, Louise Lai Sai, Thandi Sihoyiya, Paolo Denti, Rachel PJ Lai, Thomas Crede, Jonathan Naude, Patryk Szymanski, Yakoob Vallie, Ismail Abbas Banderker, Muhammed S Moosa, Peter Raubenheimer, Sally Candy, Curtis Offiah, Gerda Wahl, Isak Vorster, Gary Maartens, John Black, Graeme Meintjes, Robert J Wilkinson

## Abstract

**Background:** Drug regimens which include intensified antibiotics alongside effective anti-inflammatory therapies may improve outcomes in Tuberculous Meningitis (TBM). Safety data on their use in combination and in the context of HIV is needed to inform clinical trial design.

**Methods:** We conducted a phase 2 open-label parallel-design RCT to assess safety of high-dose rifampicin, linezolid and aspirin in HIV-associated TBM. Participants were randomised (1.4:1:1) to three treatment arms (arm 1, standard of care (SOC); arm 2 SOC + additional rifampicin (up to 35mg/kg/day)) + linezolid 1200mg/day reducing after 28/7 to 600mg/day; arm 3, as per arm 2 + aspirin 1000mg/day) for 56 days, when the primary outcome of adverse events of special interest (AESI) *or* death was assessed.

**Results:** 52 participants were randomised. 59% had mild disease (MRC Grade 1) vs 39% (Grade 2) vs 2% (Grade 3). 33% of participants had microbiologically-confirmed TBM; vs 41% ‘possible’ or 25% ‘probable’. AESI or death occurred in 10/16 (arm 3) vs 4/14 (arm 2) vs 6/20 (arm 1) (*p=0*.*083*). The cumulative proportion of AESI or death (Kaplan-Meier method) demonstrated worse outcomes in arm 3 vs arm 1 (*p=0*.*04*), however only one event in arm 3 was attributable to aspirin and was mild. There was no difference in efficacy (Modified Rankin Scale) at day 56 between the three arms.

**Conclusions:** High-dose rifampicin and adjunctive linezolid can safely be added to SOC in HIV-associated TBM. Larger studies are required to evaluate whether potential toxicity associated with these interventions, particularly aspirin, is outweighed by mortality or morbidity benefit.

**SUMMARY:** In this phase 2a randomised control trial we demonstrate that high-dose rifampicin and adjunctive linezolid is safe in adult HIV-associated tuberculous meningitis. Larger studies are required to evaluate potential toxicity with aspirin, in relation to benefit on morbidity and mortality.

## INTRODUCTION

Tuberculous meningitis (TBM) is the most severe form of tuberculosis. With currently available treatment mortality is high; up to 50% in those co-infected with HIV^1^. In those who survive, there is a high burden of disability due to neurological sequelae such as stroke^2^, epilepsy^3^, inflammatory complications within the spinal cord^4^ and neurocognitive impairment^5^. Drug regimens used to treat TBM are largely based on those used in pulmonary tuberculosis. There is a need to design and evaluate new regimens that account for the differing ability of drugs to penetrate the central nervous system (CNS) whilst simultaneously counteracting the dysregulated immune response which occurs in TBM in order to improve outcomes in this disease.

Rifampicin at standard adult doses seldom achieves adequate cerebrospinal fluid (CSF) concentrations ^6^; it is therefore likely that higher doses would increase bactericidal activity within the CNS. Two randomised controlled trials (RCT) in TBM have evaluated high-dose rifampicin (13mg/kg intravenous^7^ and 15mg/kg oral^8^) with conflicting results. A recent pharmacokinetic (PK) study suggested that doses higher than 15mg/kg may improve outcomes by demonstrating ∼8 and ∼6 fold higher CSF exposures with 35mg/kg (oral) and 20mg/kg (IV) doses respectively compared to standard oral dose (10mg/kg)^9^. Linezolid, which is now part of the WHO recommended treatment in drug resistant TB^10^, is known to have broad tissue penetration, including into the CNS^11^. Two observational studies of linezolid found favourable clinical and laboratory outcomes in both children and adults with TBM^12,13^. However, frequently reported haematological and neuropathic toxicity are concerning^14^. In TBM this toxicity, which is rarely severe^15^ and largely reversible^16^, may be acceptable. Aspirin targets key pathogenic processes which occur during TBM: inhibition of thromboxane and platelet aggregation^17^ at lower doses (75mg daily), and inhibition of proinflammatory prostaglandins and thromboxane A_2_ ^18^ at higher doses (>600mg daily). The latter may be further augmented by aspirin’s ability to trigger production of pro-resolving lipid mediators^19^. Three RCT have evaluated aspirin at varying doses (from 75mg daily to 1000mg/day) with varying outcomes in TBM^20-22^. The latest of these demonstrated reduction in infarcts and death with 1000mg of aspirin compared to placebo in HIV-negative patients with microbiologically confirmed TBM^20^. The safety of high-dose aspirin, which may lead to anti-inflammatory effects, has yet to be evaluated in the context of HIV and in conjunction with adjunctive antimicrobial drugs.

A number of clinical trials are investigating either the PK properties of linezolid (NCT04021121, NCT03537495) or efficacy of high-dose rifampicin (ISRCTN15668391) as single adjunctive therapies in TBM, and one phase 3 trial aims to assess efficacy of linezolid, high-dose rifampicin, and lower-dose aspirin (NCT04145258). However, safety data on the use of rifampicin and linezolid in combination, of high-dose aspirin in combination with intensified antibiotics, and in the context of HIV co-infection is absent. LASER-TBM aimed to generate much needed safety data on the use of enhanced antimicrobial therapy including higher-dose rifampicin (35mg/kg) and linezolid (1200mg reducing to 600mg daily) with or without adjunctive high-dose aspirin (1000mg daily) in HIV-associated TBM, to inform their use in definitive clinical trials.

## METHODS

LASER-TBM was an open label, parallel group, randomised, multi-arm phase 2A trial in which participants were randomised to one of three treatment arms as specified below. Investigational drugs were given for the first 56 days of therapy. Primary endpoint data was collected at day 56. Treatment was subsequently continued per South African national guidance. Participants completed study follow up at 6 months. Interim analysis for safety was performed by an independent data safety monitoring board (DSMB) board after every 15 participants enrolled. A full version of the study protocol is published elsewhere^23^.

### Study participants and sites

Adults aged 18 or over with a diagnosis of possible, probable or definite TBM as per uniform case definition^24^, and a confirmed HIV-1 seropositivity were eligible for enrolment. Exclusion criteria are listed in Box S1 (supplementary appendix). Written informed consent was obtained from participants where individuals were assessed to have capacity; in those without capacity to consent, proxy consent from next of kin was obtained. In the latter cases deferred consent was obtained from the patient as soon as they were able.Potential participants were referred whilst inpatients at four regional referral hospitals across South Africa. Subsequent follow-up occurred in inpatient wards and outpatient clinics at respective sites, or at two TB hospitals in Cape Town.

### Intervention

Participants were randomised to one of three treatment arms (1.4:1:1). Proportionally more participants were randomised to arm 1 to account for anticipated higher mortality with standard of care, compared to the intervention arms.

- Arm 1 (standard of care): rifampicin 10 mg/kg, isoniazid (H) 5 mg/kg, ethambutol (E) 15 mg/kg, and pyrazinamide (Z) 25 mg/kg daily for 56 days
- Arm 2: as per arm 1 plus adjunctive 25mg/kg rifampicin (total dose 35mg/kg) and linezolid (1200mg for 28 days, reducing to 600mg for 28 days) daily for 56 days
- Arm 3: as per arm 2 plus adjunctive aspirin (1000mg) daily for 56 days

Dosing was calculated by weight bands as published in the study protocol^23^. After 56 days participants were referred to government TB facilities to receive continuation therapy (rifampicin 10mg/kg/day and isoniazid 5mg/kg/day) for 7 months as per national South African guidelines^25^.

Participants allocated to arms 2 and 3 were further randomised (1:1) to receive oral rifampicin 35 mg/kg or IV rifampicin 20 mg/kg once daily for the first 3 days of therapy (in addition to HZE and linezolid with or without aspirin, according to the experimental arm). Results of this PK sub-study are published elsewhere^26^.

### Outcome Measures

The primary endpoint of the study was the cumulative proportion of participants experiencing AESI or dying by 56 days. AESI were selected based on anticipated toxicity related to the two interventional arms: bleeding (gastrointestinal bleeding and intracerebral haemorrhage); haematological (anaemia, neutropenia, thrombocytopenia); transaminitis; and neuropathic (peripheral and optic neuropathy) (see Table 1). Secondary endpoints included death and severe disability by 56 days, death by 56 days and 180 days; disability at 56 and 180 days; incidence of grade 3 or 4 adverse events, permanent discontinuation of study drugs, severity and frequency of haematological and neurological AESI related to linezolid use; severity and frequency of major bleeding (gastrointestinal and intracerebral) related to aspirin use; occurrence of TBM-Immune Reconstitution Inflammatory Syndrome (IRIS) assessed by the modified International Network for the Study of HIV-associated IRIS (INSHI) criteria^27^; and MRI and CT changes at day 56.

**Table 1:**
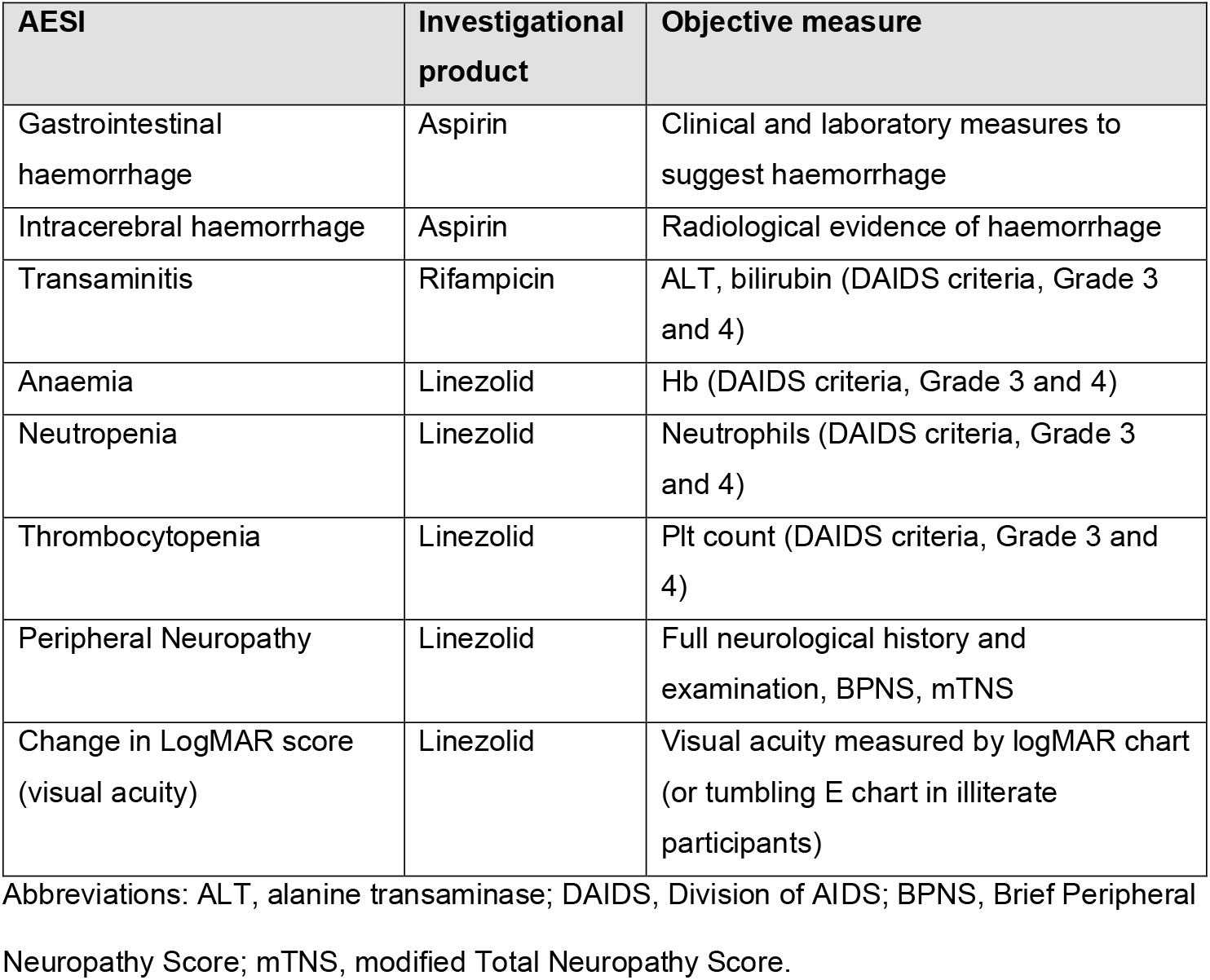
Adverse events of special interest (AESI) assessed in the trial.

| AESI | Investigational product | Objective measure |
| --- | --- | --- |
| Gastrointestinal haemorrhage | Aspirin | Clinical and laboratory measures to suggest haemorrhage |
| Intracerebral haemorrhage | Aspirin | Radiological evidence of haemorrhage |
| Transaminitis | Rifampicin | ALT, bilirubin (DAIDS criteria, Grade 3 and 4) |
| Anaemia | Linezolid | Hb (DAIDS criteria, Grade 3 and 4) |
| Neutropenia | Linezolid | Neutrophils (DAIDS criteria, Grade 3 and 4) |
| Thrombocytopenia | Linezolid | Plt count (DAIDS criteria, Grade 3 and 4) |
| Peripheral Neuropathy | Linezolid | Full neurological history and examination, BPNS, mTNS |
| Change in LogMAR score (visual acuity) | Linezolid | Visual acuity measured by logMAR chart (or tumbling E chart in illiterate participants) |

Abbreviations: ALT, alanine transaminase; DAIDS, Division of AIDS; BPNS, Brief Peripheral Neuropathy Score; mTNS, modified Total Neuropathy Score.

### Study schedule and safety outcome measures

Participants were screened and enrolled within 5 days of commencing TB treatment. They were assessed at five subsequent study visits (days 3, 7, 14, 28, 56) at which point they were referred to government TB clinics for continuation phase therapy (Figure S1).

Participants were assessed clinically at one further timepoint (day 180) either in person or telephonically. Where feasible, imaging (either CT and/or MRI) was performed at baseline and day 56. A neurological assessment including full motor and sensory examination of the limbs and cranial nerve examination was performed at each study visit. Participants’ functional neurological status was assessed using the Modified Rankin Scale at each study visit up to day 56 and telephonically at day 180. Assessment for AESI was also made at each study visit up to and including day 56 involving; a clinical history to assess symptoms of bleeding (GI or intracerebral), BPNS and modified total neuropathy score to assess for peripheral neuropathy, a LogMAR or tumbling E charts to assess for optic neuropathy, a full blood count to assess for haematological abnormalities and liver function tests to assess for a transaminitis. A full list of clinical outcome measures and assessments performed at each study visit are listed in the published protocol^23^.

### Study oversight

An independent DSMB oversaw the safety of the trial and advised to continue recruitment without change after review of primary endpoint data for each 15 participants enrolled. No formal stopping rules were stipulated but rather were left to the discretion of the DSMB.

Approval for the trial was granted by the University of Cape Town Human Research Ethics Committee (293/2018), Walter Sisulu University Human Research Committee (012/2019) and the South African Health Products Regulatory Authority (20180622). The trial was registered on the South African National Clinical Trials Register (DOH-27-0319-6230), Pan African National Clinical Trials Register (PACTR201902921101705) and clinicaltrials.gov (NCT03927313).

## STATISTICAL ANALYSIS

### Sample size

No formal statistical power calculation was performed. Even as single adjunctive therapies there was limited available data on the use of these drugs in TBM to predict likely rate of AESI and/or death. Given this would be further complicated when considering likely event rate when the drugs were combined, it was felt a more pragmatic approach was to create a recruitment target of 100 participants with frequent blinded review of cumulative safety events by an independent DSMB. An aim of LASER-TBM was also to serve as a planning study to generate PK and safety data to inform a phase 3 RCT of intensified treatment in TBM (NCT04145258), which in part would influence resulting sample size of that study. The rate of recruitment was slower than anticipated due to the COVID-19 pandemic and in January 2021 a decision was made, in consultation with the DSMB, to cease recruitment to allow commencement of the aforementioned phase 3 RCT.

### Statistical analysis

Analysis was performed in GraphPad Prism v.9.0 and R v.3.6.0. The primary analysis was performed in the modified intent-to-treat population (those who receive any dose of the study drug). A sensitivity analysis was planned for the per-protocol population (those who completed treatment as specified in the protocol) however given the small sample size and since these populations were similar, here we report the most conservative analysis (modified intention to treat).

The primary endpoint, frequency of AESI or death (where data is censored at the first event prior to day 56) was summarized and compared across arms using a chi-squared test. A time to event analysis was performed for worst grade (in each individual participant) AESI or death; comparisons between study groups were made using the log-rank test. Neurological disability (measured by Modified Rankin Score), as well as radiological outcomes at day 56 were compared across treatment arms using chi-squared test. We used spaghetti plots to visually represent longitudinal CSF parameters (lymphocytes, polymorphonuclear cells, protein and glucose) over time and used t tests to compare longitudinal summaries (mean and SD) of each individual trajectory across treatment arms.

Details of further analysis can be found in the full statistical analysis plan published alongside the study protocol^23^.

## RESULTS

98 patients were screened and 52 were randomised between June 2019 and January 2021 (Figure 1). Reasons for screening exclusion are summarised in Table S1. One participant was randomised but excluded prior to any study IP being dispensed due to emergence of an exclusion criterion (eGFR <20) on a hospital blood test performed prior to randomisation. Another participant was excluded from the modified intention to treat analysis as they died prior to receiving any dose of study drug. Six participants discontinued the study prior to day 56, and a further four participants discontinued between day 56 and day 180 (Table S2).

**Figure 1:**
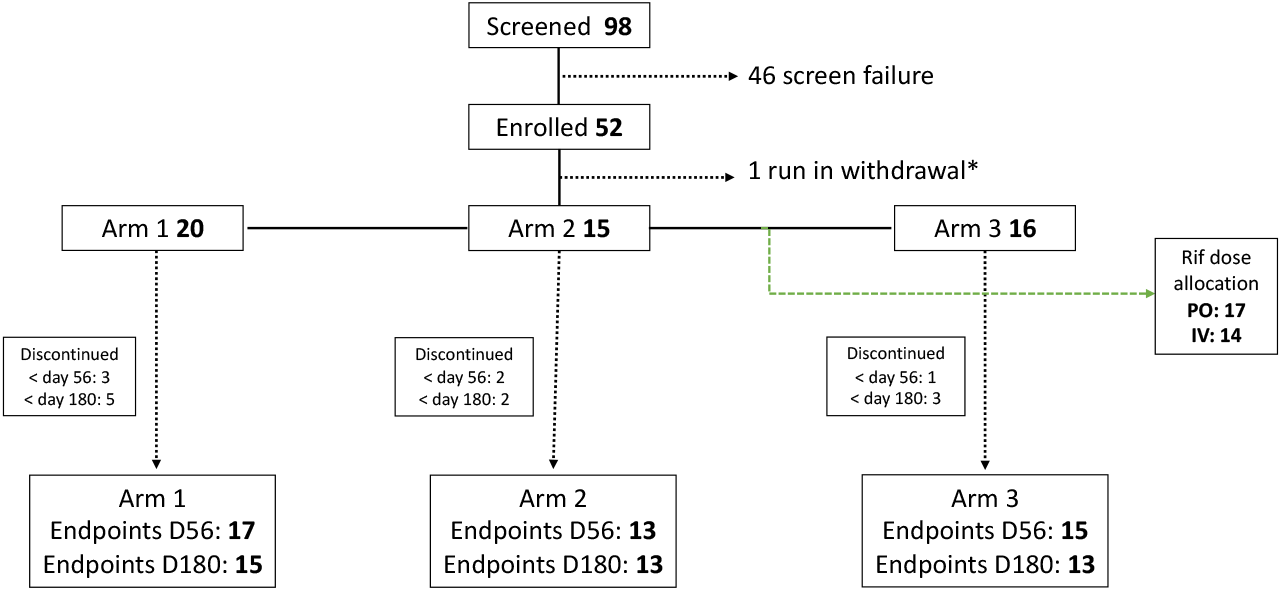
Consolidated Standards of Reporting Trials (CONSORT) diagram. Abbreviations: Rif, rifampicin *patient randomized but withdrawn prior to receiving study IP due to emergence of exclusion criteria Legend: CONSORT diagram to describe recruitment and arm allocation. Reasons for screening exclusions and early study withdrawals are listed in Tables S1 and S2

The baseline characteristics of the participants stratified by treatment arm are described in Table 1. Most participants were male (71%) and the median age was 39 (34-46). Most participants had mild disease (MRC Grade 1 59%; Grade 2 39%; Grade 3 2%). A third (33%) of participants had microbiologically confirmed TBM at baseline, with the remaining participants defined as possible (41%) or probable TBM (25%) as per the uniform TBM case definition^24^.

The primary endpoint analysis was performed in the modified intention to treat population (n=50; arm 1, 20; arm 2, 14; arm 3, 16). The composite primary endpoint of AESI or death occurred in 6/20 in arm 1, 4/14 in arm 2, and 10/16 participants in arm 3, (p=0.083). The occurrence of each category of AESI stratified by treatment arm are summarised in table 3 with further detail on timing and outcome of each of these events listed in table 4. Frequency of death prior to day 56 was similar across arms (n=7; arm 1, 3; arm 2, 1; arm 3, 3; *p=0*.*649*) and in no case was cause of death related to study investigational product (table 5). Grade 3 or 4 AE (grade 3: arm 1, 7 vs arm 2, 7 vs ar, 3, 9, *p=0*.*44*; grade 4: arm 1, 2 vs arm 2, 4 vs arm 3, 4, *p=0*.*38*) or serious adverse events for reasons other than death (arm 1, 6 vs arm 2, 8 vs arm 3, 7, *p=0*.*37*) were similar across treatment arms.

**Table 2:** Baseline characteristics.

|  | Arm 1 (n=20) | Arm 2 (n=15) | Arm 3 (n=16) |
| --- | --- | --- | --- |
| Age, median (IQR) years | 39.5 (34-48.5) | 37 (34.5–42.5) | 41.5 (31.8-46) |
| Gender, male n (%) | 10 (50) | 10 (66.7) | 16 (62.5) |
| Uniform case definition, n (%) |  |  |  |
| - Definite | 8 (40) | 3 (20) | 6 (37.5) |
| - Probable | 5 (25) | 4 (26.7) | 4 (25.0) |
| - Possible | 7 (35) | 8 (53.3) | 6 (37.5) |
| BMRC TBM Grade, n (%) |  |  |  |
| - Grade 1 | 11 (55) | 11 (73.3) | 8 (50) |
| - Grade 2 | 8 (40) | 4 (26.7) | 8 (50) |
| - Grade 3 | 1 (5) | 0 (0) | 0 (0) |
| CD4 T-cell count, median (IQR), cells/μL | 116.5 (58.6-283) | 131 (82.5-186) | 158.5 (85.5-331.5) |
| HIV viral load, median (IQR), copies/mL | 89,150 (1000-203,711) | 37,960 (2428 – 394,839) | 2686 (1361-777,620) |
| ART status, n (%) |  |  |  |
| - On ART | 6 (30) | 5 (33) | 5 (31) |
| - Previous ART | 3 (15) | 6 (40) | 5 (31) |
| - ART naive | 11 (55) | 4 (27) | 6 (38) |
| Of those on ART, duration in weeks, median (range) | 288.9 (22.4-459.3) | 23.7 (0.4-83.6) | 355 (2.9-879.1) |
| CSF cell count/biochemical data available (n) | 17 | 14 | 13 |
| Polymorphonuclear cells, median (IQR), cells/ μL | 13 (0-85) | 4 (2-16) | 16 (3-22) |
| Lymphocytes, median (IQR), cells/ μL | 63 (10-259) | 79 (11-218) | 82 (28-278) |
| Protein, mg/dL | 1.78 (1.13-3.13) | 1.89 (0.95-4.2) | 1.9 (1.32-2.99) |
| CSF Glucose, mg/dL | 2.2 (0.9-2.5) | 2.4 (1.9-2.9) | 1.7 (1.2-3.3) |
| Baseline radiology available (n) | 16 | 12 | 11 |
| Hydrocephalus n (%) | 1 (6.3) | 1 (8.3) | 1 (9.1) |
| Meningeal enhancement n, (%) | 4 (25) | 2 (16.7) | 6 (54.5) |
| Tuberculoma(s) n, (%) | 1 (6.3) | 2 (16.7) | 2 (18.2) |
| Infarct(s) n. (%) | 4 (25) | 1 (8.3) | 3 (27.3) |

Abbreviations: IQR, interquartile range; ART, antiretroviral therapy; BMRC, British Medical Research

**Table 3:** AESI by treatment arm.

|  | **Arm 1**<br>**(n=20)** | **Arm 2**<br>**(n=14)** | **Arm 3**<br>**(n=16)** | ***p value* \*\*** |
| --- | --- | --- | --- | --- |
| Bleeding, n* (%) | 0 (0) | 0 (0) | 1 (6) | 0.338 |
| Transaminitis, n (%) | 0 (0) | 0 (0) | 2 (13) | 0.109 |
| Hematological, n (%) | 2 (10) | 0 (0) | 1 (6) | 0.481 |
| Peripheral Neuropathy, n (%) | 2 (10) | 2 (14) | 4 (25) | 0.46 |
| Change in LogMAR score, n (%) | 0 (0) | 2 (14) | 2 (13) | 0.231 |

\*individuals with an event
\*\* arm 3 vs arm 1

**Table 4:**
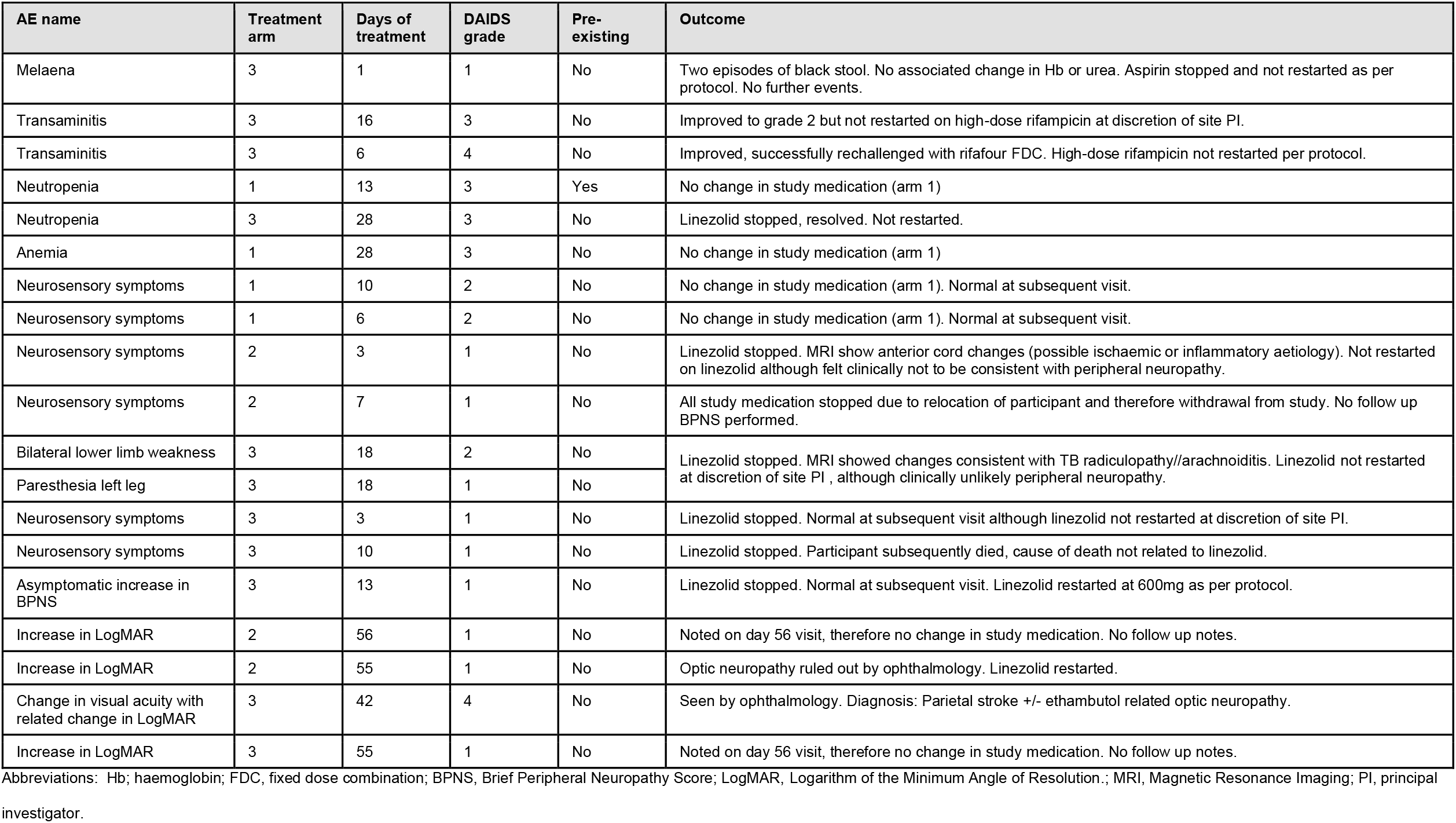
Details of AESI by event.

**Table 5:** Timing and cause of death prior to day 56.

| Cause of death | Treatment Arm | Days of IP |
| --- | --- | --- |
| Renal Failure | 1 | 11 |
| TB Meningitis | 1 | 15 |
| TB Meningitis | 1 | 2 |
| TB Meningitis | 2 | 8 |
| TB Meningitis | 2 | 0* |
| Pulmonary embolism | 3 | 39 |
| TB Meningitis | 3 | 6 |
| TB Meningitis | 3 | 3 |

\*death prior to receiving study IP and therefore excluded from modified intention to treat population analysis

The cumulative incidence of the composite endpoint of worst grade AESI or death at day 56 demonstrated worse outcomes when comparing arm 3 vs arm 1 (p=0.043), with similar proportions observed in other pre-specified analysis (arm 2 *vs* arm 1 (*p=0*.*3*), arm 2+3 combined *vs* arm 1 (*p=0*.*5*)) (Figure 2, log rank test). Similarly, analysis for death alone demonstrated no difference between arms (Figure 3). The cumulative incidence of AESI events was greater in arm 3 vs arm 1 (p=0.02), however, when arms 2 and 3 were combined and compared to arm 1 this difference was less marked (p=0.18) (Figure 4).

**Figure 2:**
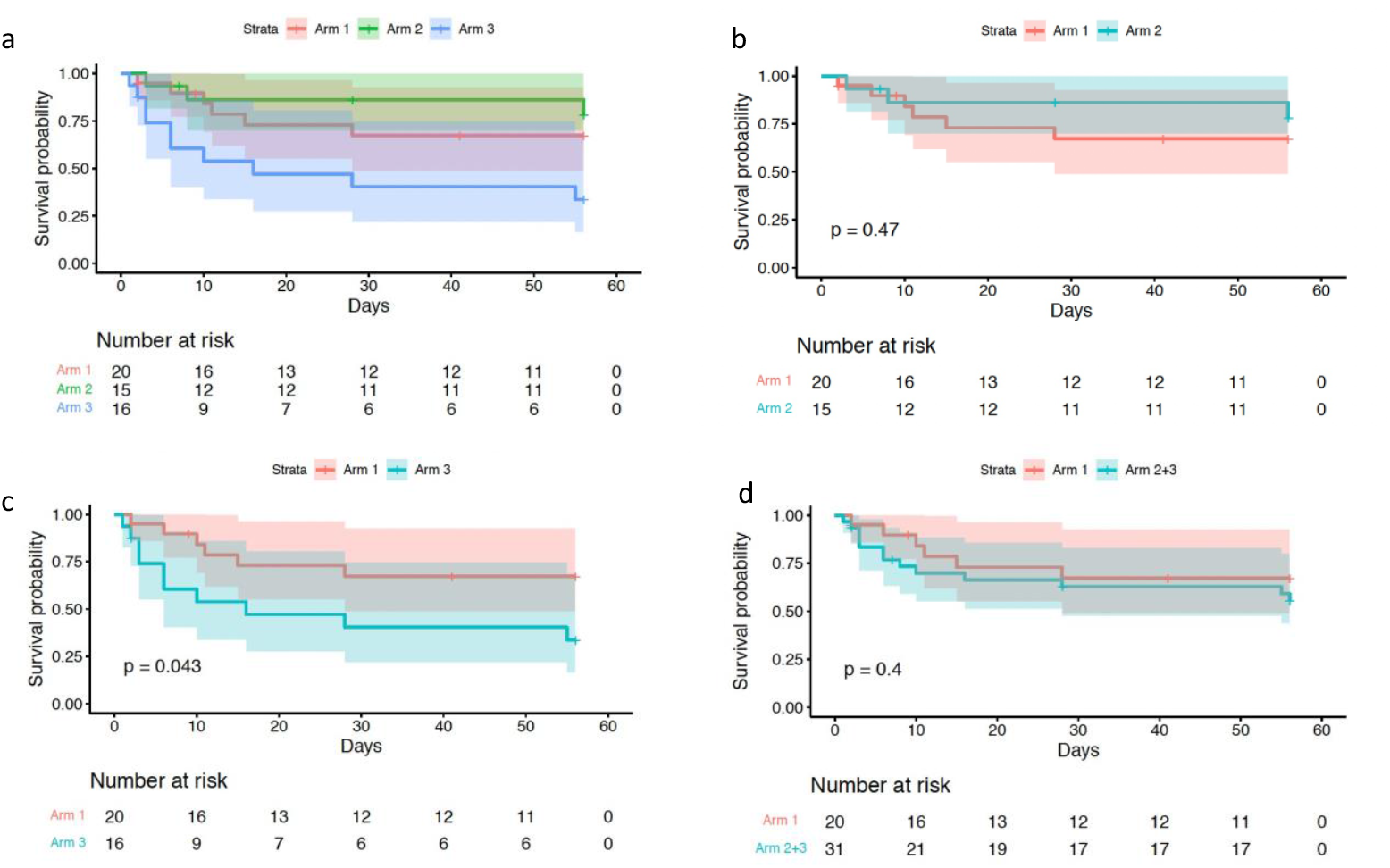
Time to worst grade AESI or death. Kaplan-Meier analysis of time to worse grade AESI or death, comparing arm 1, 2 and 3 (a), arm 2 vs arm 1 (b), arm 3 vs arm 1 (c) and arm 2 and 3 combined vs arm 1 (d).

**Figure 3.**
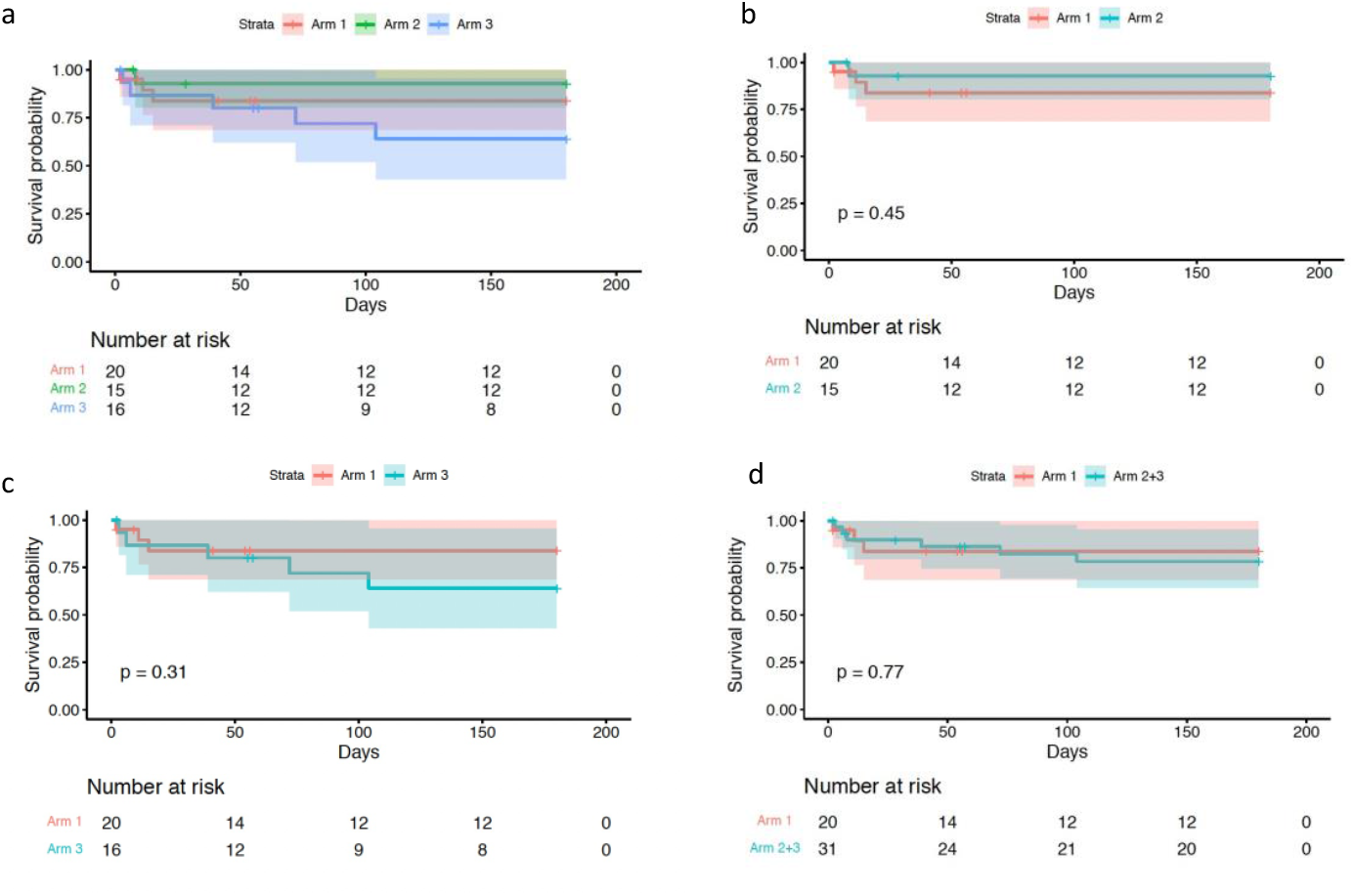
Time to death. Kaplan-Meier analysis of time to death, comparing arm 1, 2 and 3 (a), arm 2 vs arm 1 (b), arm 3 vs arm 1 (c) and arm 2 and 3 combined vs arm 1 (d).

**Figure 4.**
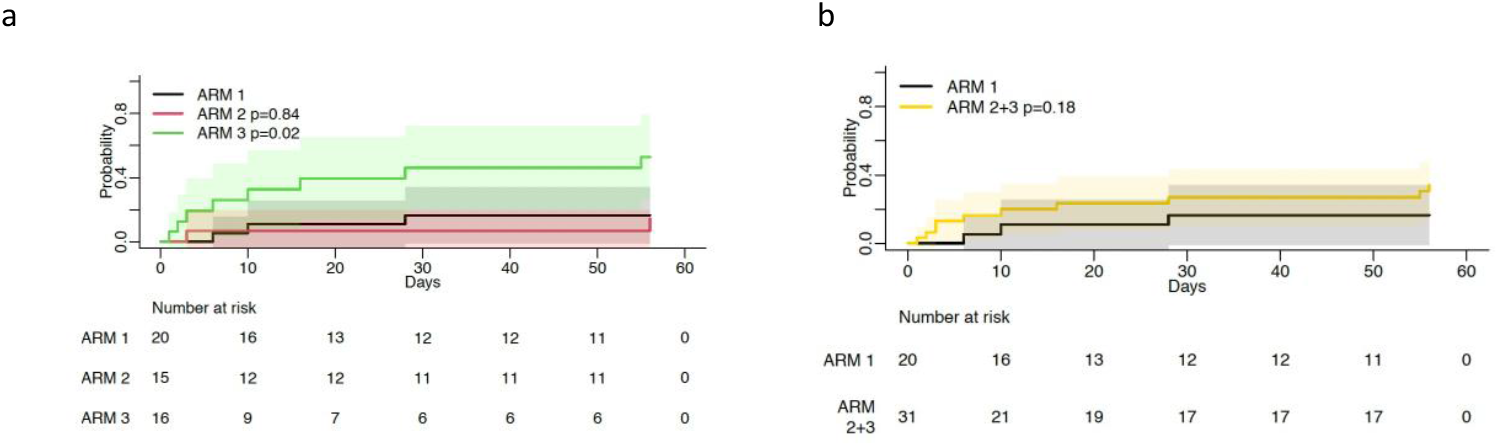
Time to AESI. Kaplan-Meier analysis of time to AESI, comparing arm 1, 2 and 3 (a), and arm 2 and 3 combined vs arm 1 (b).

The frequency of grade 5 MRS (severe disability) *or* death was 4 (arm 1) vs 3 (arm 2) vs 5 (arm 3), p=0.774. The frequency of good (defined as MRS grade 0-3), and bad outcomes (MRS Grade 4-6) were similar across arms (p=0.616) (Figure 5). *Post hoc* analysis of change in neurological function (as measured by MRS) found similar changes of MRS from baseline to day 56 between the three arms (Figure 4a). Few IRIS events occurred (arm 1, 2; arm 2, 2; arm 3, 3), of which 4/7 were defined as neurological IRIS. Within the first 56 days of treatment, four participants developed new onset lower limb weakness (TB myelopathy 2; TB radiculomyelopathy/arachnoiditis 1; other (no cause found prior to death) 1); three participants developed a new onset hemiplegia; one patient developed a new onset isolated cranial nerve palsy (lower motor neuron VII). Thirteen participants presented with new onset seizures at TBM diagnosis. A further nine participants had new onset seizures within the first 2 months of follow up (arm 1, 5; arm 2; 2; arm 3, 2; *p=0*.*54*). Baseline and follow up imaging was performed in only 9 patients at the timepoints pre-specified within the protocol. Follow up imaging demonstrated new or worsening leptomeningeal enhancement in 2/9 participants (arm 1 and arm 2), new evidence of infarction in 2/9 participants (arm 1 and arm 2), new or worsening tuberculomas in 2/9 participants (arm 1 and arm 2) which was associated with worsening sulcal effacement in 1/9 participant (arm 1).

**Figure 5:**
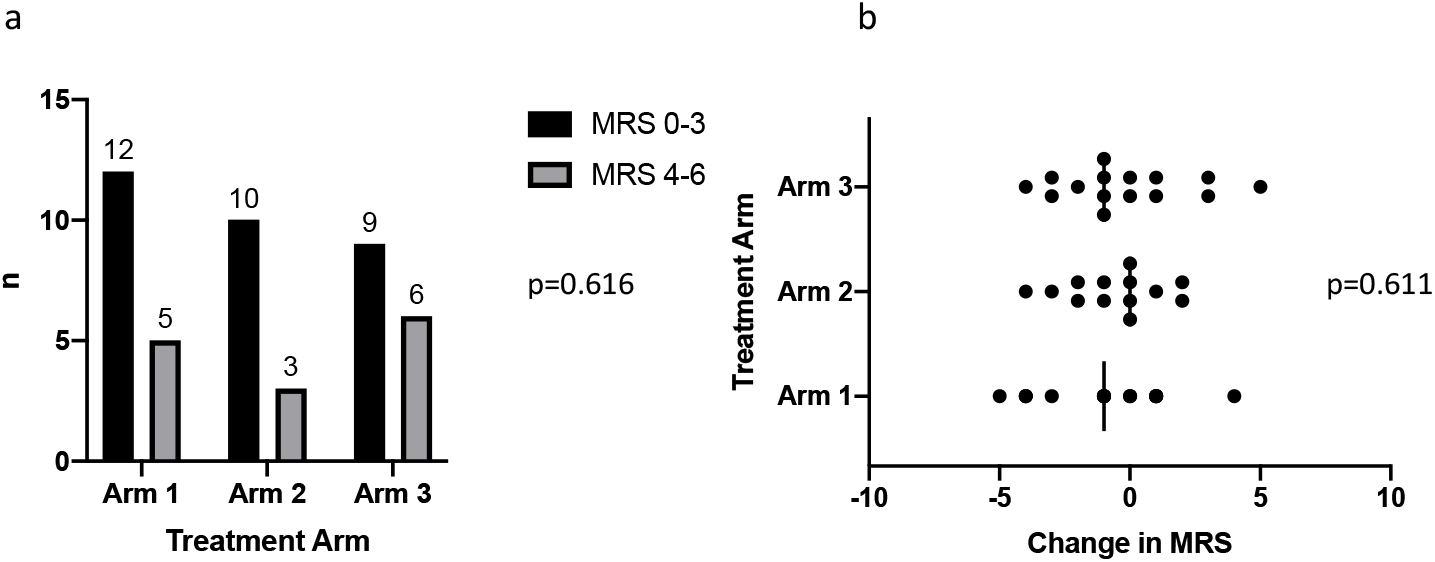
Functional outcome at day 56 as defined by Modified Rankin Scale. Figure 3a demonstrates a comparison between good outcome (MRS 0-3) vs bad outcome stratified by arm at day 56. Figure 3b compares change in MRS between enrolment and day 56, across treatment arms.

Spaghetti plot analysis of longitudinal CSF parameters (lymphocyte and polymorphonuclear cell count, protein and glucose) over time demonstrated downward trend of parameters across all three treatment arms (figure 6). Individual values are plotted and the superimposed line represents the mean values at each timepoint in each treatment arm. T tests comparing mean and variance at each time point demonstrated no difference between arms.

**Figure 6:**
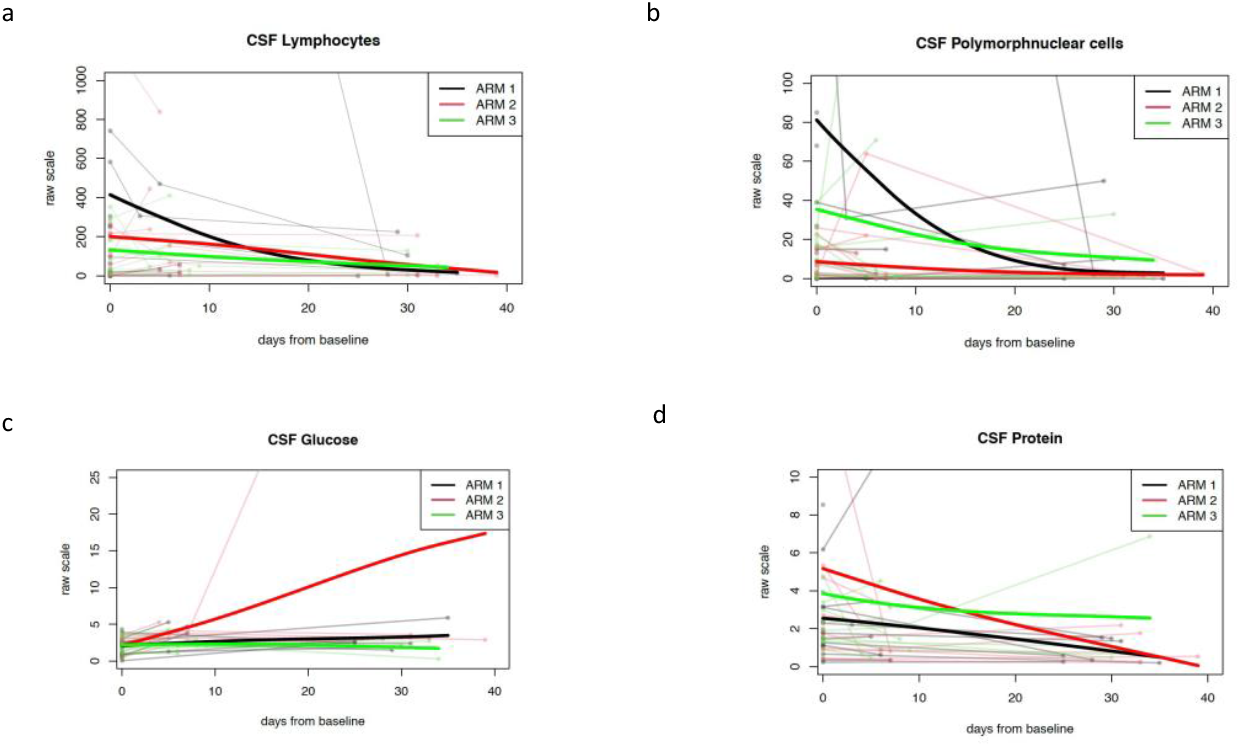
Change in CSF parameters over time. Spaghetti plots for CSF parameters (lymphocyte count (a), polymorphonuclear cells (b), glucose (c), protein (d)) plotted as individual values over time (faint lines), with mean values for each treatment arm represented by superimposed line (bold lines).

## DISCUSSION

The LASER-TBM study was a phase 2a RCT which evaluated the safety of high-dose rifampicin (35mg/kg daily), adjunctive linezolid (1200mg reducing to 600mg after 28 days) and adjunctive aspirin (1000mg daily) for the first 56 days of treatment in HIV-associated TBM. Primary endpoint analysis showed no significant difference in the incidence of AESI or death between treatment arms, although there was a trend towards an increase in events in arm 3. There was no difference in death or disability at day 56 across arms; and a similar frequency of clinical or radiological events occurred in each arm. Exploratory analysis found no difference in change in CSF parameters over time by arm.

Although secondary analysis revealed a significantly higher number of events (AESI or death) in arm 3 vs arm 1 (*p=0*.*04*), it is reassuring that no deaths were attributed to aspirin. Only one bleeding event occurred, after minimal exposure to high-dose aspirin (1 dose), resolved immediately following discontinuation of the drug, and was not associated with any laboratory markers to suggest significant gastrointestinal bleeding. Toxicity attributable to linezolid was similarly mild; of seven events potentially attributable to linezolid occuring in participants randomised to experimental arms, 3/7 were due to an alternative cause and 2/7 had recovered prior to the subsequent study visit. No patient was formally diagnosed with peripheral neuropathy, which is expected given most recent studies showing a median time to onset of neuropathy occurs after 10 weeks of treatment^15,16^. Only one participant developed a change in visual acuity, which may have been due to linezolid, although on review by an ophthalmologist was assessed more likely due to ethambutol. The number of participants in whom potential abnormalities were detected using the LogMAR and tumbling E assessments, compared to the confirmed number of cases of optic neuropathy calls into question the specificity of these outcome measures. Given that linezolid has potential to treat TBM as well as drug resistant TB we must consider whether better outcome measures can be developed to reliably detect abnormalities attributable to these drugs in both the clinical and research setting in order to prevent overestimation of toxicity, particularly when used for a short duration. Toxicity due to rifampicin was similarly infrequent with only two participants developing clinically significant transaminitis. In both cases the transaminitis recovered with treatment interruption. These results suggest that toxicity associated with the enhanced antitubercular regimen (rifampicin 35mg/kg and adjunctive linezolid) is not common when used in combination for two months to treat HIV-associated TBM. This is encouraging in the context of a disease where no specific evidenced-based antitubercular regimen exists, and provides rationale for the ongoing phase 3 RCT (NCT04145258) where participants are randomised to receive both high-dose rifampicin and linezolid at doses identical to that used in this study.

There are several limitations to this study. Although no formal power calculation took place, final sample size was substantially smaller than the target of 100 participants. The COVID-19 pandemic adversely affected recruitment to the study, and in January 2021 a decision was made to stop recruitment to enable commencement of a similar phase 3 study which was ready to start resulting in a lower than proposed sample size. It is unknown whether the significantly higher number of AESI or death in arm 3 vs arm 1 demonstrated within the secondary analysis reflects a true safety risk of the regimen containing aspirin, or is due to chance given the lower than anticipated numbers of participants recruited. Secondly, the majority participants recruited had mild TBM. The reasons for this is likely multifactorial including i) patients dying prior to screening given that up to 5 days of TB treatment was allowed prior to enrolment and ii) patients with decreased levels of consciousness arriving at hospital alone and therefore not having available next of kin available for proxy consent. In the latter case a protocol amendment was approved to allow deferred consent in these patients, however, is likely to explain in part the higher rate of mild disease in our cohort. The mild level of disease within our patient cohort likely explains the low level of mortality; 16% 2-month mortality contrasted the oft quoted 50% mortality within the literature^1^. The primary endpoint of AESI *or* death was designed with the assumption that observed mortality would be near or approaching 50%. The relatively few numbers of deaths led to a greater proportion of AESI in the composite endpoint of AESI *or* death. Given that all of the listed AESI were proportionally more likely to occur in the experimental arm 3, it is unsurprising that the number of events within the composite endpoint of AESI or death occurred in arm 3 where the greatest number of interventions was given. This is supported by the observation that when considering AESI alone, the cumulative incidence of events was significantly greater in arm 3, suggesting that the composite endpoint of AESI or death was driven by the higher rate of AESI in arm 3.

Ours is the first RCT to evaluate linezolid in TBM, an important drug in this context. It is the first completed trial to focus on an exclusively HIV-positive population, a vulnerable group in TBM. It is also the first study to date to systematically evaluate the safety of a novel drug regimen containing enhanced antitubercular treatments alongside a host directed therapy in TBM. To improve outcomes in TBM therapeutic advance must simultaneously aim to enhance bacterial clearance within the CNS which controlling the dysregulated host inflammatory response. Our data provides rationale for the safe use of high-dose rifampicin and linezolid in TBM and supports ongoing evaluation of the efficacy of these drugs in a phase 3 trial. It is less clear whether high-dose aspirin is safe in this context. Given the results of previous studies which did not demonstrate any safety concerns, as well as its potential to target key pathways which play a role in the pathogenesis of TBM a larger study is now required to see if potential harm is offset by a morbidity and mortality benefit.

## Data Availability

All data produced in the present work are contained in the manuscript

## AKNOWLEDGMENTS

We would like to thank firstly the participants and their carers for agreeing to take part in the study. We would like to acknowledge staff at referring hospitals, at Western Cape and Eastern Cape provincial governments, and at UCT Clinical Research Centre for their help with making the study a success. We would like to thank the Data Safety and Management Board for their advice and guidance throughout the trial, namely; Professor David Lalloo (chair, Liverpool School for Tropical Medicine), Dr David Meya (College of Health Sciences, Makerere University and Division of Infectious Diseases and International Medicine, University of Minnesota), Dr Evelyne Kestelyn (Clinical Trials Unit, Centre for Tropical Medicine and Global Health, University of Oxford), Dr Maryline Bonnet (Institute of Research for Development, France), Dr Angela Crook, (MRC Clinical Trials Unit, University College London).

## SUPPLEMENTARY MATERIAL

**Figure S1:**
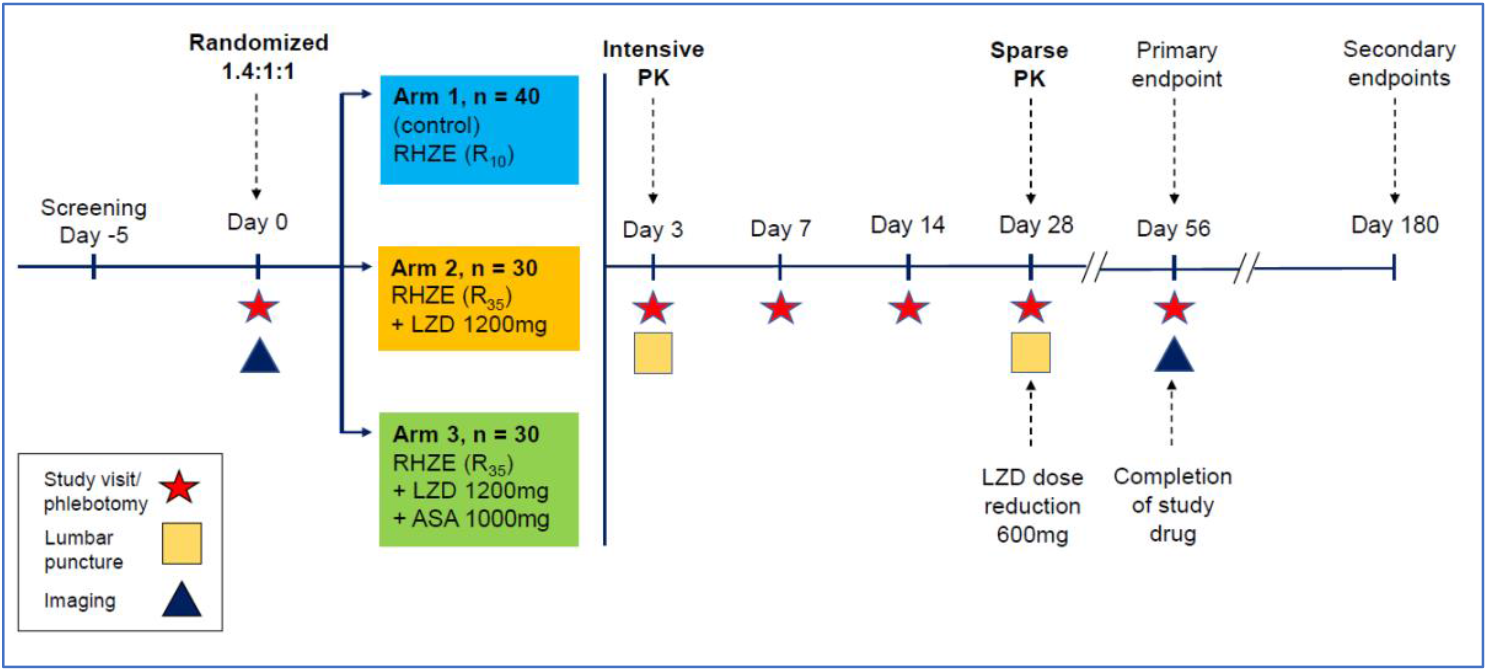
Study schedule. Legend: Study design schematic describing randomisation to study arms, treatment intervention per am, visit schedule, overview of clinical procedures and timepoints relating to primary and secondary endpoint data collection Abbreviations: RHZE: Rifampicin, Isoniazid, Pyrazinamide, Ethambutol; R_10_: Rifampicin 10mg/kg/day; R_35_: Rifampicin 35mg/kg/day; LZD: Linezolid; ASA; Aspirin;

**Table S1:**
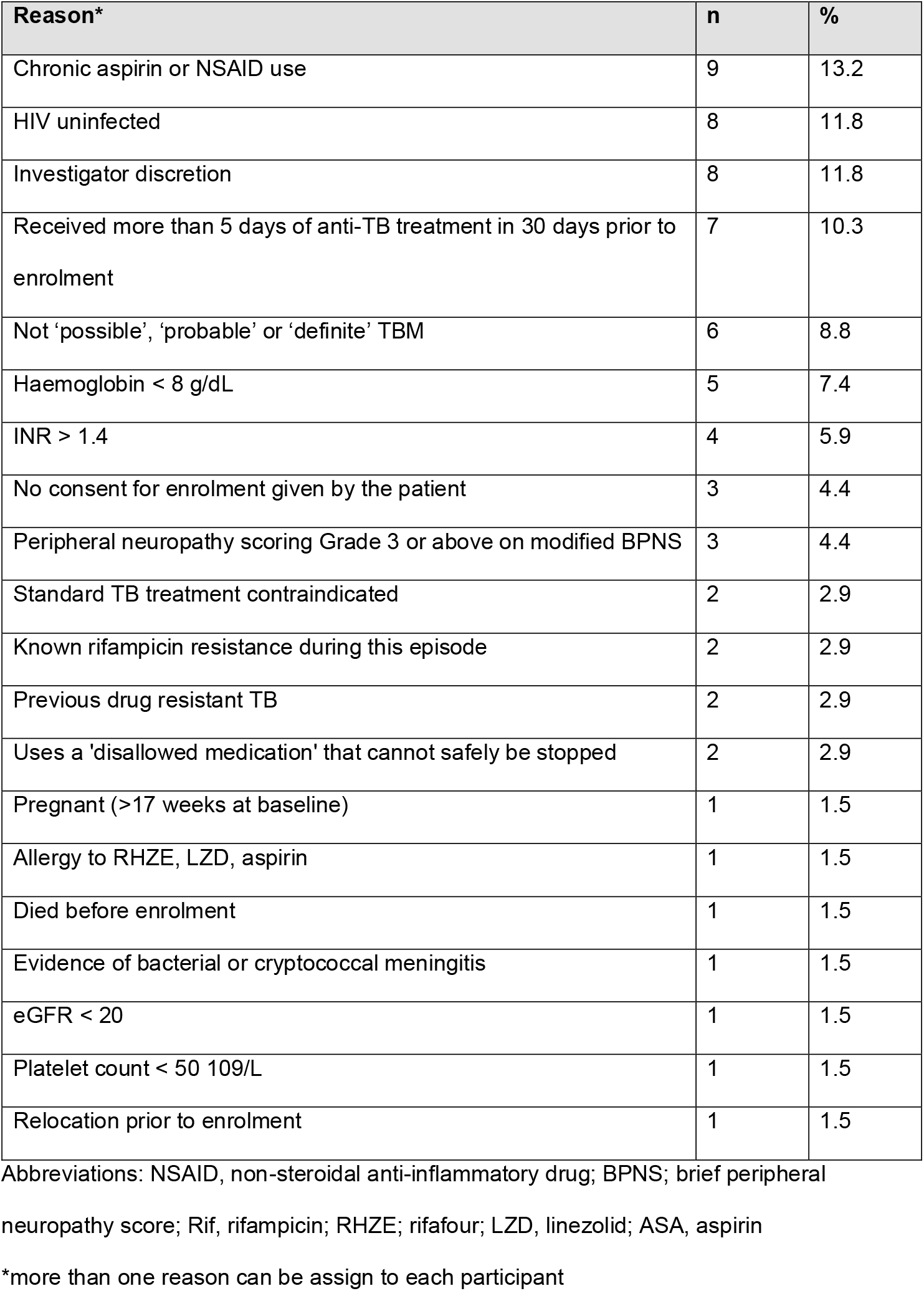
Reasons for screening exclusion.

| Reason* | n | % |
| --- | --- | --- |
| Chronic aspirin or NSAID use | 9 | 13.2 |
| HIV uninfected | 8 | 11.8 |
| Investigator discretion | 8 | 11.8 |
| Received more than 5 days of anti-TB treatment in 30 days prior to enrolment | 7 | 10.3 |
| Not 'possible', 'probable' or 'definite' TBM | 6 | 8.8 |
| Haemoglobin < 8 g/dL | 5 | 7.4 |
| INR > 1.4 | 4 | 5.9 |
| No consent for enrolment given by the patient | 3 | 4.4 |
| Peripheral neuropathy scoring Grade 3 or above on modified BPNS | 3 | 4.4 |
| Standard TB treatment contraindicated | 2 | 2.9 |
| Known rifampicin resistance during this episode | 2 | 2.9 |
| Previous drug resistant TB | 2 | 2.9 |
| Uses a 'disallowed medication' that cannot safely be stopped | 2 | 2.9 |
| Pregnant (>17 weeks at baseline) | 1 | 1.5 |
| Allergy to RHZE, LZD, aspirin | 1 | 1.5 |
| Died before enrolment | 1 | 1.5 |
| Evidence of bacterial or cryptococcal meningitis | 1 | 1.5 |
| eGFR < 20 | 1 | 1.5 |
| Platelet count < 50 10<sup>9</sup>/L | 1 | 1.5 |
| Relocation prior to enrolment | 1 | 1.5 |

Abbreviations: NSAID, non-steroidal anti-inflammatory drug; BPNS; brief peripheral
neuropathy score; Rif, rifampicin; RHZE; rifamur; LZD, linezolid; ASA, aspirin
\*more than one reason can be assign to each participant

**Table S2:** Reason for study withdrawals prior to day 56.

| Withdrawal | Reason |
| --- | --- |
| 1 | Participant relocated to Malawi and therefore unable to attend study follow up visits |
| 2 | Participant relocated to Kwazulu-Natal and therefore unable to attend study follow up visits |
| 3 | Participant withdrew consent |
| 4 | Participant withdrew consent |
| 5 | Participant developed acute psychosis and was unsafe to follow up |
| 6 | Participant lost to follow up |

### Box S1

Eligibility Criteria

#### Inclusion criteria

- Age >18 years
- proven HIV-1 seropositivity
- Diagnosis of ‘possible’, ‘probable’ or ‘definite’ TBM

#### Exclusion criteria

- Rifampicin-resistant *M*.*tb* detected on any clinical specimen;
- History of allergy or hypersensitivity to RIF, isoniazid, ethambutol, pyrazinamide, LZD or ASA;
- Received more than 5 days of antitubercular therapy in the 30 days prior to screening;
- Receipt of regular daily ASA or NSAID prior to TBM diagnosis
- CSF unobtainable by lumbar puncture or another procedure;
- Evidence of bacterial or cryptococcal meningitis;
- Severe concurrent uncontrolled opportunistic infection including, but not limited to, active cytomegalovirus-associated disease, Kaposi sarcoma, *Pneumocystis jirovecii* pneumonia, HIV related or unrelated malignancy, or gastrointestinal bleeding;
- Any other form of immunosuppressive therapy, including antineoplastic and biologic agents, apart from corticosteroids;
- More than 17 weeks pregnant at baseline;
- Peripheral neuropathy scoring Grade 3 or above on the BPNS;
- Any disease or condition in which the use of the standard anti-TB drugs (or any of their components) are contraindicated. This includes, but is not limited to, allergy to any TB drug or their components;
- The presence of one or more of the following:
- Estimated glomerular filtration rate (eGFR) < 20ml/min/1.73 m2^*^
- INR > 1.4 and/or clinical evidence of liver failure or decompensated cirrhosis;
- Haemoglobin < 8.0 g/dL;
- Platelets < 50 ×109 /L;
- Neutrophils < 0.5 × 109 cells/L;
- Any disease or condition in which any of the medicinal products listed in the section pertaining to prohibited medication is used and cannot be safely stopped;
- Known or suspected history of drug abuse or any other reason that is, in the opinion of investigators, sufficient to compromise the safety or cooperation of the participant.

## Footnotes

* Calculated using the Cockcroft-Gault equation; INR: International normalised ration; BPNS: Brief Peripheral Neuropathy Score; NSAID: Non Steroidal Anti Inflammatory Drug;

